# How can rural community-engaged health services planning affect sustainable health care system changes? - A process description and qualitative analysis of data from the Rural Coordination Centre of British Columbia’s Rural Site Visits Project

**DOI:** 10.1101/2020.11.19.20232769

**Authors:** C Stuart Johnston, Erika Belanger, Krystal Wong, David Snadden

## Abstract

**Objectives:** The objectives of the Rural Site Visit Project (SV Project) were to develop a successful model for engaging all 201 communities in rural British Columbia, Canada, build relationships and gather data about community health care issues to help modify existing rural health care programs and inform government rural health care policy.

**Design:** An adapted version of Boelen’s health partnership model was used to identify each community’s Health Care Partners: health providers, academics, policy makers, health managers, and community representatives. Qualitative data was gathered using a semi-structured interview guide. Major themes were identified through content analysis, and this information was fed back to the government and interviewees in reports every six months.

**Setting:** The 107 communities visited thus far have health care services that range from hospitals with surgical programs to remote communities with no medical services at all. The majority have access to local primary care.

**Participants:** Participants were recruited from the Health Care Partner groups identified above using purposeful and snowball sampling.

**Primary and secondary outcome measures:** A successful process was developed to engage rural communities in identifying their health care priorities, whilst simultaneously building and strengthening relationships. The qualitative data was analysed from 185 meetings in 80 communities and shared with policy makers at governmental and community levels.

**Results:** 36 themes have been identified and three overarching themes that interconnect all the interviews, namely Relationships, Autonomy and Change Over Time, are discussed.

**Conclusion:** The SV Project appears to be unique in that it is physician led, prioritizes relationships, engages all of the health care partners singly and jointly in each community, is ongoing, provides feedback to both the policy makers and all interviewees on a 6-monthly basis and, by virtue of its large scope, has the ability to produce interim reports that have helped support system change.

**Article Summary:**

- This study process has adapted Boelen’s health partnership model and is unique in that it is physician led, prioritizes relationships, engages all of the health care partners singly and jointly in each community, is ongoing, provides feedback to both the policy makers and all interviewees on a 6-monthly basis.
- A successful method of engaging with rural communities and building relationships and trust across multiple stakeholder groups is described that contributed to influencing positive health care system changes.
- As all communities in one province are being visited a picture of rural health care initiatives and challenges is highly comprehensive and therefore able to influence policy.
- One of the main limitations in this study is that because the interviewers were experienced health care providers, power differentials may have existed which may have introduced bias in the discussions.
- A potential limitation is the enormous amount of data to handle and analyze in a rigorous way, which was mitigated by having two full time analysts working together to ensure consistency with frequent meeting with the research team to consider and agree emerging themes.

How can rural community-engaged health services planning affect sustainable health care system changes? – A process description and qualitative analysis of data from the Rural Coordination Centre of British Columbia’s Rural Site Visits Project

## Introduction

British Columba (BC), Canada, has a population of approximately 5 million. About fourteen percent (631,776) (1) are rural citizens distributed unevenly over an area of 944,738 km^2^. BC is geographically diverse with a broken 27,000 km coastline and extensive mountain ranges that make for long and often dangerous travel, complicated at times by wildfires, floods, avalanches and harsh winter conditions. Access to health care services for rural citizens is often limited by the expansive geography, provider availability (2) and transportation issues (3).

Support programs for rural physicians in BC are overseen by the Joint Standing Committee on Rural Issues (JSC), a committee comprised of equal numbers of provincial Ministry of Health representatives and rural physicians. The JSC manages approximately C$150M (2020) of funding annually for programs and projects that improve health care delivery in rural BC (https://www2.gov.bc.ca/assets/gov/health/practitioner-pro/rural-guide.pdf). Some of this work is delivered by the Rural Coordination Centre of BC (RCCbc), which is funded by the JSC to coordinate and improve rural health care throughout the province.

The Rural Site Visits Project (SV Project) was initiated in 2017 by rural physicians with a proposal to the JSC who tasked the RCCbc with visiting 201 rural and Indigenous BC communities identified as eligible for rural benefits under the RSA https://www2.gov.bc.ca/gov/content/health/practitioner-professional-resources/physician-compensation/rural-practice-programs/rural-practice-subsidiary-agreement. The RSA is an agreement between the Government of BC and the Doctors of BC (a professional organization that represents 14,000 physicians, medical residents and medical students in BC. https://www.doctorsofbc.ca/).

The purpose of the SV Project was to build relationships between rural physicians, health care providers, health administrators, municipal leadership, First Nations leadership, first responders, academia and policy makers through listening and gathering data systematically about local successes, innovations and challenges relating to rural health care delivery. This data is guiding the development of JSC programs and informing government Rural Health Care policy.

In 1978 the declaration of the Alma-Ata International Conference on Primary Health Care stated that: “The people have the right and duty to participate individually and collectively in the planning and implementation of their health care” (4). Current trends in rural health services, however, aim to reduce infrastructure and support to achieve greater efficiencies through centralization of services (5, 6). Small rural communities have had to be proactive in securing local health services to resist this development (7, 8), requiring improved relationships and communication between the policy makers and communities.

Community participation has been seen as a more complete approach to health development (9) leading to culturally and contextually appropriate decisions being made about rural health services (10, 11). Relationship building between stakeholders is also seen as more effective than attempting to provide a myriad of health care services (12, 13), especially as each rural community is unique and “one size fits all” approaches are largely ineffective (6, 14). While there have been efforts by health service policy makers to align their actions with rural communities’ expressed priorities (15, 16), the processes used for community engagement have received less attention (17) and descriptions seldom include adequate documentation of the processes involved (17, 18).

The community engagement literature does not show examples of rural health projects initiated and led by physicians, even though physicians have been key partners in other research on rural community-engaged health services planning (15). Much of the research on community engagement in rural health service planning has had a specific focus, for example in improving immunization programs in Nigeria (17) or chronic disease care in the Torres Strait Islands (13). There are some examples of research focused on community participation for broader primary care reform, for example, in the Northern Health Authority region of BC (15) and the Remote Service Futures (RSF) Project in Scotland (10, 12, 16). The former has resulted in some sustained changes to date, for example the establishment of Primary Care Nurses, improved antenatal care and regional palliative care services (15). When the RSF outcomes were reviewed in 2014: “Only one direct sustained service change was found” (19). These raise the question of how best to affect sustainable beneficial rural health system changes using community engagement processes. The project described here attempts to address this issue.

## Design & Methods

### Theoretical Approach

The Health Partnership model described by Boelen (20) was used. This identifies five partners: health professionals, academic institutions, policy makers, health managers and citizens and recommends they meet to identify ways to improve health systems. The concept of meeting with the partners together in each community was modified to include additional separate meetings with each of the partners. Who constituted the health partners could be different in each community, so the concept was adapted to the local context to include those present in the community. This could include others such as first responders, business and non-profit groups. It was not possible to have combined partner meetings in all communities as it was not always possible to find a date and time within the visits time line that worked for everyone.

The interviews incorporated an Appreciative Inquiry approach (21, 22) with intentional listening using a semi-structured interview guide. The interviews focused on how rural community members perceived health care delivery within their respective communities seeking successes and innovations as well as challenges. To process the large volume of qualitative data collected, qualitative content analysis (23) was used.

### Patient and Public Involvement

Public input into the research project occurred during the initial pilot Site Visits to eight rural communities.

Public input was used to shape the community engagement process and the interview guide.

The initial interview guide was developed by the investigators, who had many years of rural health care experience, to elicit broad discussion about multiple health care issues. The guide was refined based on public and provider input during pilot visits. The interview format continued to be iteratively improved based on feedback from subsequent Site Visits.

Persons representing the health care partner groups in each community were recruited initially. Snowball recruitment was then used to include other valuable perspectives.

Participants were asked for feedback on the interview process and whether the time taken was appropriate.

Every six months a Community Feedback Report is circulated to all past interviewees in which the latest results are discussed. The report is in the public domain and dissemination is encouraged.

### Site Recruitment

The sites identified for the SV Project were the 201 communities identified under the RSA.

### Arranging Site Visits

Sites are selected six to twelve months in advance. Three to six months prior to a Site Visit, recruitment of participants commences and RCCbc staff coordinate the planning. Depending on community size and location site visits last one to three days and involve one to five communities.

### Site Visits Team

A Site Visits team consists of at least one Site Visitor and one RCCbc staff member, who coordinates the visits. The Site Visitors comprise 19 rural physicians and one midwife. A one-day training session for interviewers included training in Appreciative Inquiry techniques and qualitative interviewing and cultural safety through the San’yas Indigenous Cultural Safety Course (http://www.sanyas.ca/). Site Visitors were individually mentored by the Program Leads on their first visits. On some Site Visits guests are invited. The purpose of inviting a guest is to assist urban-based allies in their understanding of how health care functions in small rural communities. Guests have included policy makers, researchers, health care workers, administrators, and educators.

### Participant Recruitment

The study population included participants who identified themselves as living or working in an RSA community and were part of one or more of the partner groups identified by Boelen (20). Participants were recruited using purposeful and snowball sampling (23) through the following methods:

- Email and phone contact through publicly available information
- Recruitment posters in doctors’ lounges, hospitals, clinics, and municipal buildings
- Contacting pre-existing contacts who provide connections to potential participants
- Asking participants to suggest others who fit the inclusion criteria

Initial contact was made by telephone or e-mail with a follow-up invitation that detailed the project background, aims and goals and included a copy of the interview guide and consent form. Participants were invited to participate in one-on-one interviews or focus groups (if there was more than one person from an identified health partner group) and dates established. Interviews took place in the communities, however since March 2020, eleven virtual interviews have been trialed as a result of Covid-19 restrictions.

### Data Collection

Each health partner group (between one and sixteen participants) was interviewed separately. This was followed by a combined partner focus group (between two to ten people) with a representative from each of the health partner groups previously interviewed. A semi-structured interview guide was used which has been iteratively refined following community visits, in keeping with standard qualitative methods (supplementary file “Inteview Guide”). The guide was informed by Appreciative Inquiry and public input in order to build relationships and to better understand how rural community members perceive health care delivery within their respective communities including health care successes, innovations and challenges that inhibit their ability to access services in an equitable manner. Interviews were recorded digitally and transcribed. Interviews generally lasted one hour. Transcripts were returned to participants within four weeks for verification, alteration, or withdrawal if requested.

### Data Analysis

NVivo 12 (QSR International) was used to help organize the data. Initially each interview was coded using an inductive-approach and primary cycle coding (23). This began with a close reading of the data, assigning words or phrases that captured the essence of each sentence. From this a codebook was developed (supplementary file “Code Book”), and second level codes were generated to identify emerging themes across the data. Throughout analysis data was revisited to allow for the comparison and modification of codes to fit new incoming data.

Rigor was maintained throughout by a second data analyst. Analysts coded identical interviews separately and then compared coding to promote consistency. Analysts met weekly to discuss changes and modifications needed for the coding framework. The coding framework and emerging analysis was discussed and agreed within the research team. The data was further interpreted to identify themes connecting the data across communities (23).

### Knowledge Translation

Emerging themes are disseminated to policy makers, physicians, allied health professionals, First Nations, municipality members, academics, and the general public through various knowledge translation outputs such as a six-monthly JSC and publicly available community feedback reports (https://ruralsitevisits.rccbc.ca/project-documentation/)and newsletters, specialized (focused) reports, presentations, briefing notes, and publications. Additionally, an inovations website (https://ruralsitevisits.rccbc.ca/innovations/) has been established to share successful innovations identified by interviewees.

## Ethics

The study received harmonized ethics approval from the Behavioral Research Ethics Board of the University of British Columbia. Operational approval was also received from each health authority. Informed consent is collected from all participants.

## Results

### Site Visits Engagement Process

Although the Covid-19 pandemic has significantly slowed down the project, 382 interviews have been carried out in 107 communities over a three-year period (Table 1). The first 4 site visits to 9 communities with 23 interviews were used to pilot and develop the methods and were not included in the analysis reported here which is based on 185 interviews with 754 participants in 80 communities. The data from the remaining 27 site visits are being transcribed, returned to participants and analysed. As the data is well saturated and the processes take several months it seems appropriate to report the study now.

**Table 1.**
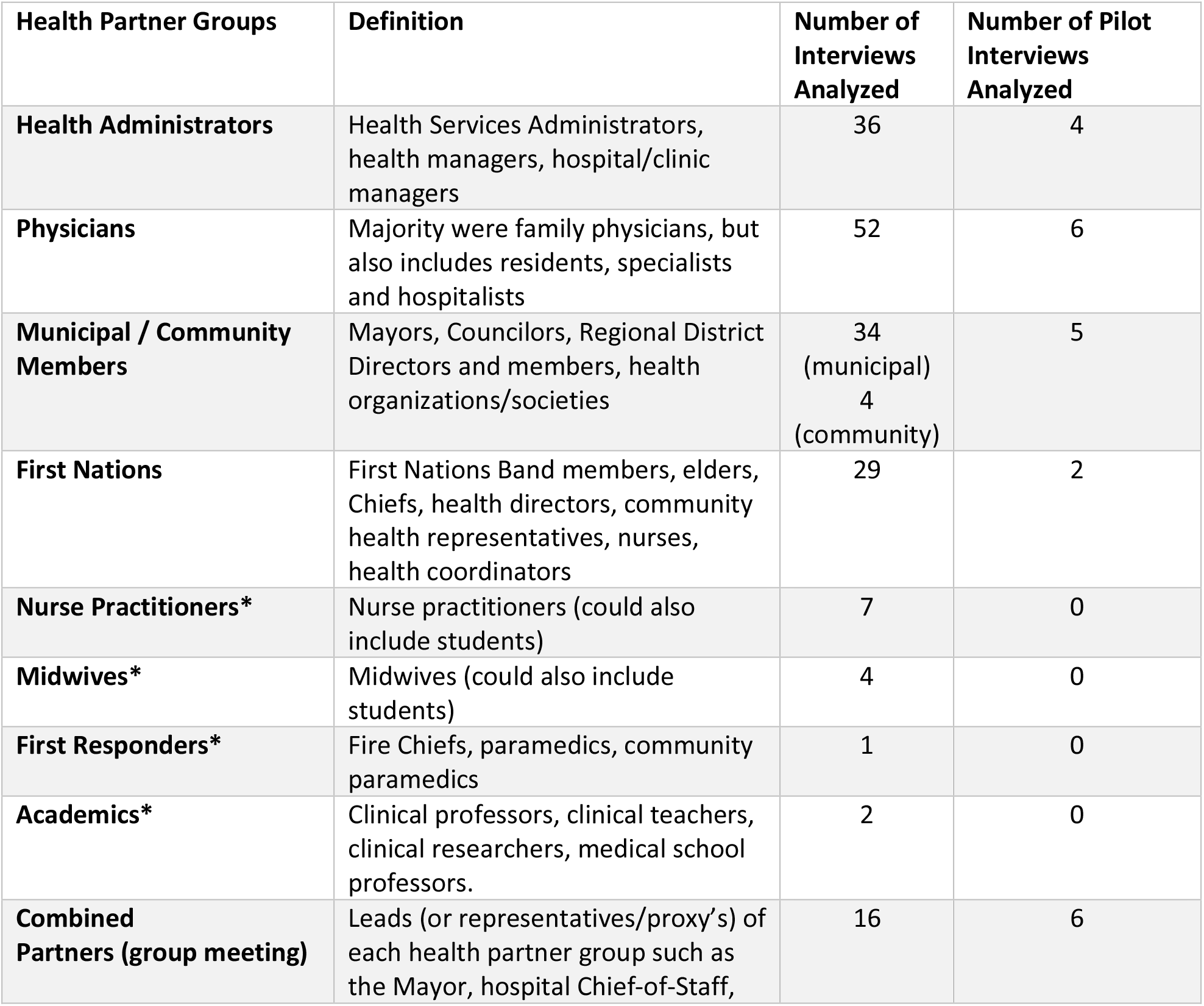
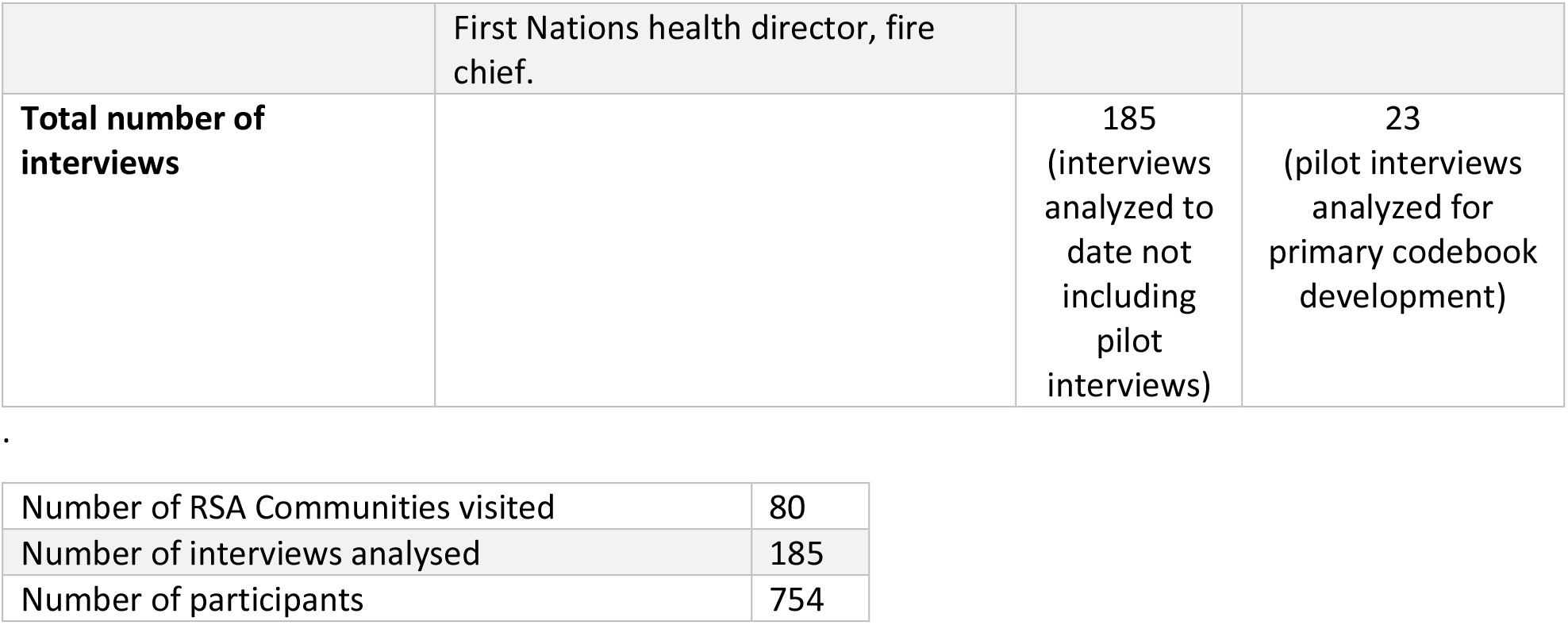
Partner groups and numbers of interviews

| Health Partner Groups | Definition | Number of Interviews Analyzed | Number of Pilot Interviews Analyzed |
| --- | --- | --- | --- |
| **Health Administrators** | Health Services Administrators, health managers, hospital/clinic managers | 36 | 4 |
| **Physicians** | Majority were family physicians, but also includes residents, specialists and hospitalists | 52 | 6 |
| **Municipal / Community Members** | Mayors, Councilors, Regional District Directors and members, health organizations/societies | 34<br>(municipal)<br>4<br>(community) | 5 |
| **First Nations** | First Nations Band members, elders, Chiefs, health directors, community health representatives, nurses, health coordinators | 29 | 2 |
| **Nurse Practitioners\*** | Nurse practitioners (could also include students) | 7 | 0 |
| **Midwives\*** | Midwives (could also include students) | 4 | 0 |
| **First Responders\*** | Fire Chiefs, paramedics, community paramedics | 1 | 0 |
| **Academics\*** | Clinical professors, clinical teachers, clinical researchers, medical school professors. | 2 | 0 |
| **Combined Partners (group meeting)** | Leads (or representatives/proxy's) of each health partner group such as the Mayor, hospital Chief-of-Staff, | 16 | 6 |
|  | First Nations health director, fire chief. |  |  |
| **Total number of interviews** |  | 185<br>(interviews analyzed to date not including pilot interviews) | 23<br>(pilot interviews analyzed for primary codebook development) |

|  |  |
| --- | --- |
| Number of RSA Communities visited | 80 |
| Number of interviews analysed | 185 |
| Number of participants | 754 |

Across interviews collectively, one participant withdrew their transcript. Many participants provided feedback; highlighting their enjoyment of the direct, in-person engagement process that was used and the connections they provided:

> *“I think this has been very informative. Just getting to know what you guys do*…*and* [*the*] *supports* [*that exist*] *and establishing connections and*…*learning about these connections that exist that I haven’t tapped into personally so, it’s great*.*” – Combined Partners*

Participants further described how they felt the process allowed for their voices to be heard, and their communities to be recognized:

> *“I appreciate being able to talk*…*and to give frank feedback because that is tough at times and this is a good option to do it*…*some of our issues aren’t really out there right? So, it’s good to be able to have a voice to be able to indicate this*.*” – Nurse Practitioner*
>
> *“I want to thank you for recognizing us a ‘rural,’ because a lot of people don’t see us as rural*.*”*
>
> *– First Nations*

It was commonly voiced by participants that, throughout the engagement process, they’d love to learn about what other communities have achieved.

> *“Would love to see information about other initiatives going on around other provinces that they might be able to learn from*.*” – Combined Partners*
>
> *“*[*We*] *would like to receive feedback about how* [*we*] *work with other communities and what works well in other communities*.*” – Combined Partners*

These requests from participants ultimately led to the creation of the Site Visits Innovations web site

### Site Visits Themes

The data has become well saturated with 36 categories emerging from the data to date. The ten most common themes are presented briefly to provide context (Table 2), and these will be the subject of subsequent publications. This article reports three overarching themes that interconnect all the data: Relationships, Autonomy and Change Over Time.

**Table 2:**
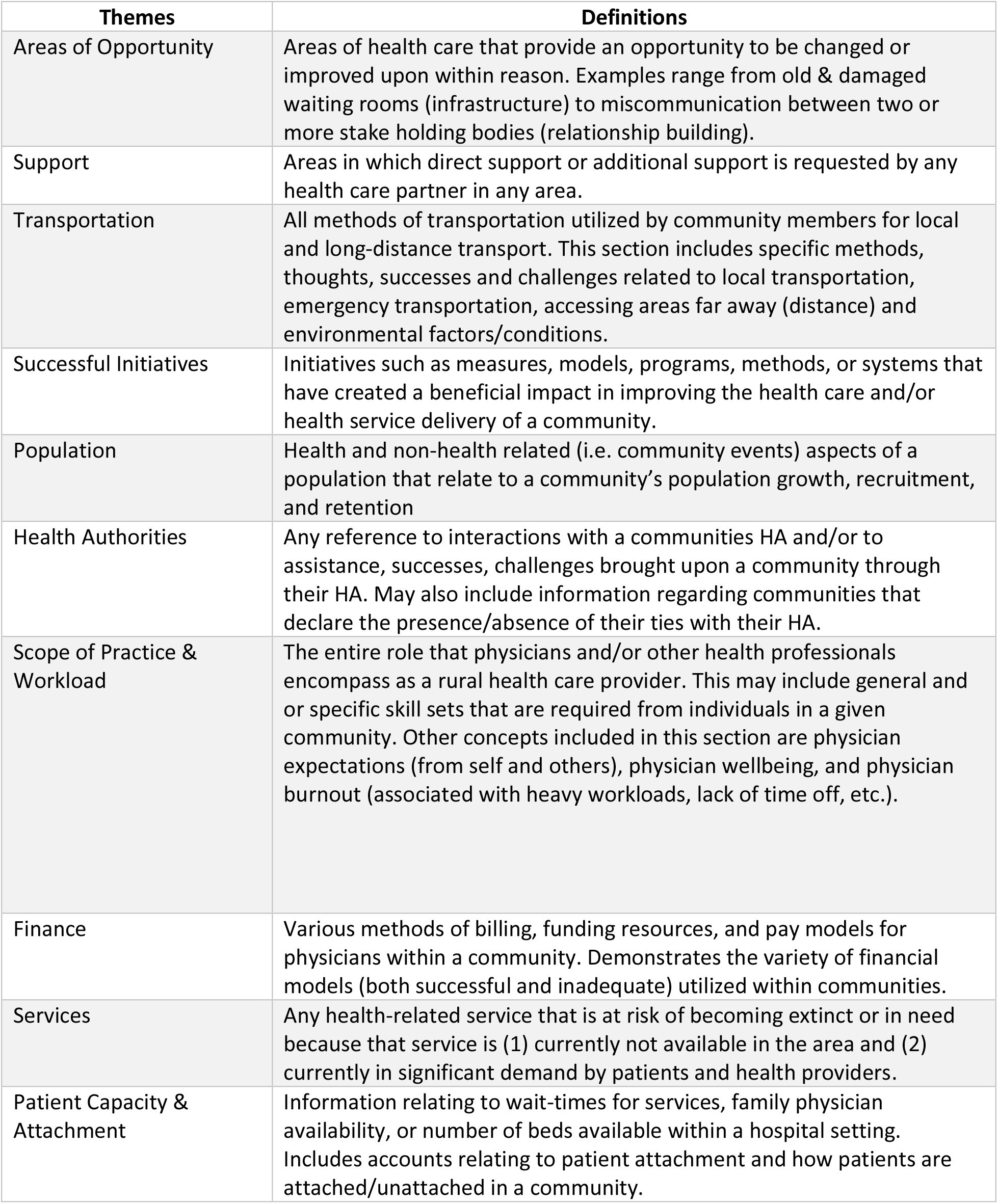
Rural Site Visits Project Table 2: List of Top 10 Themes

#### Relationships

Relationships were important in achieving successful health care outcomes and were built on communication, trust, transparency and collaboration over time. These themes were evident in every community:

> *“It’s really groups of people coming together on committees that have people from city council, the regional district, health boards, and the non-profit societies*…*and I think if there’s a strength in this community, it’s that there are those connections and people are willing to work together to find solutions locally*.*” – Combined Partners*

Good relationships underpinned communities’ abilities to successfully retain their physicians. These relationships were with the communities themselves as well as with administrators and within teams:

> *“Why do you think they’ve stayed here?” – Interviewer*
>
> *“*[*It’s*] *the relationship that they* [*the physicians*] *maintain with the community*…*It all comes down to the relationships*.*” – Municipality*
>
> *“When we went to* [*Health Authority X*] *to say, ‘We’re having a terrible time retaining our doctors,’ - the turnover was terrible - we got no response from the system. So, the community rallied around and did what was necessary to sustain doctors in this community. But in doing that* …. *what we did was create relationships with our physicians that are respectful and goes* [*both*] *ways*.*” – Community Members*

Effective communication and regular “organic” contact were the foundation of these relationships and were important in building trust:

> *“Having all the different services all in the one building does allow for good open communication, you can pull anyone aside if you bump into them in the hallway to talk about patients. It is a very organic process rather than a formalized team-based care approach. It also helps retain people who work here – you build that relationship and trust of what your peers are capable of. It’s not formal team-based care, but it is a team*.*” – Combined Partners*
>
> *“There needs to be trust and consistency of knowing what someone is walking into. Issues of trust* [*have been*] *a major block in* [*our*] *community to providing and receiving health services*.*” – Health Admin*

Successful collaborations that were inclusive of all partners positively impacted health care and helped reduce burnout:

> *“It makes it much easier working* [*here*], *because I’ve worked here a really long time with* [*colleagues X and Y*] *it makes it much easier when we have a group that all works together really well. And that doesn’t happen everywhere*. [*We*] *are all friends so* [*we*] *tend to help each other out. being* [*without them*] …*the burnout would be terrible*.*” – Physicians*
>
> *“It’s really groups of people coming together on committees that have people from city council, the regional district, health boards, and the non-profit societies that identify the problems and look at what each particular group*…*can provide to try to deal with the problem and*…*it’s those connections and people* [*who*] *are willing to work together to find solutions locally*.*” – Combined Partners*

Conversely, poor collaboration and relationships led to adverse consequences:

> *“*…*when I meet with my doctors, I hear one thing about what the problem is and how to solve it. And then, if I talked to nurses or midwives or allied health professionals, I hear another version of what the problem is and how we would fix it. And then I sit down with* [*Health Authority X*] *and I hear their version of what the problem is and that they are fixing it. And all those voices are never in the same room at the same time*…*” – Municipality*

Good relationships enhance problem solving, reduce the ‘red tape’ required to affect change and result in greater work satisfaction at all levels, positively affecting other issues such as recruitment, retention, and burnout. Local decision making (autonomy) was an important contributor to work satisfaction.

#### Autonomy

Autonomy within the health care context was defined in many ways. However, at its core many viewed autonomy as the ability to make reasonable decisions, sensitive to the local context, at a personal or local level that did not require the blessings of a hierarchical, top-down system. The latter stifled initiative, innovations, and satisfaction.

A sense of autonomy within the health care providers appears to improve recruitment and retention. It imbued a sense of greater ‘ownership’ of, or responsibility for, the local services by the community practitioners:

> *“Part of it is the relationship that they maintain with the community*…*Dr* [*X*] *has come to the council and has asked for extra room to bring in more medical professionals, and the city worked with him so that he can have the space to have another professional help out his team. The main thing is working with them and letting them grow, not dictating to the doctors*.*” – Municipality*

The data described a disconnect between centrally directed processes and what was practically achievable in a community:

> *“*…*I think there’s kind of an issue sometimes with delivery of rural health care in that people actually in the trenches doing the job have a much better insight sometimes into what needs to be done and what is happening than the people making the decisions about how we’re going to deliver the health care*.*” – Physicians*

The most frequent plea was that more local engagement was needed to solve local problems and how important local autonomy was in crafting enduring solutions:

> *“I couldn’t believe that – ‘we are bringing more resources and that’s not working for you?’ What didn’t happen is there was no consultation, so it didn’t really matter if we brought more resources. It was like, ‘you didn’t ask us what our problem is, what we need and what is our reality and you’re just bringing resources and that’s not how we want this to look like*…*” – Health Admin*
>
> *“*…*locally it feels like our concerns are profoundly dismissed by the health authority, who clearly have a different idea and a different agenda”* … *“We need to be kind of at least a largely autonomous community*.*” – Physicians*

When consultation occurred a very different attitude existed among the health care providers:

> *“*…*So, we took that learning and stepped back and took one whole year to do focus group and to follow staff to understand what they’re doing, what are the challenges, the issues, to understand better the population that we serve*…*involving physicians along the way and after we’ve done all of this, we came up with another model, not really with much more budget*…*but it wasn’t about the budget anymore and we’ve presented the model to the staff in March and since then, we are implementing the new model and it’s working and people are just following along the process and I think that there’s a lot of learning about the history of the community and how we need to do things here*.*” – Health Admin*

Local autonomy meant the ability to make rapid operational decisions on the day. Many small rural communities had extraordinary stories of unbroken 24/7 emergency coverage for many years provided by the local practitioners despite being reduced to a single physician at times. Similarly, nurses in small rural hospitals frequently did additional shifts to cover gaps when their colleagues were unable to work. These providers felt a responsibility to maintain these services in their community:

> *“I had a lot of autonomy about who I could hire*…*and so I had the ability to hire locally and so I built a big pool of people who lived here who were very committed to* [*the*] *Healthcare Centre*.*” – Health Admin*

When control of these services was elevated to a higher level outside of the community, this loyalty was reduced as local autonomy was lost, contributing to Emergency Room coverage gaps and difficulty filling nursing shifts:

> *“*…*So now we have one GP who is keeping the whole system going through being on call 24 hours a day, 7 days a week. So, it’s sort of a step backwards, and I think a lot of it is just that we’ve lost the autonomy to be able to kind of say, “Well, this is what our community needs. This is how we can go about solving this problem*.*” – Physicians*
>
> *“*…*you’ve done a really innovative thing in adjusting your nursing lines*…*this is the first community we have not heard* [*about*] *nursing shortages*.*” – Interviewer*
>
> *“So, we need to start developing our rotations to make it attractive for those nurses to come*…*We’re one of the few rural sites that have full staffing now*.*” – Health Administrator*

One example of a successful model is a 3-year trial in a region where a Health Authority granted three geographically close rural communities the autonomy to determine their priorities for improving local health care, and provided funding to support these changes:

> *“We had a series of engagement events for the entire community, health care providers, public, youth at one of the high schools, our Indigenous population, and the* [*Community X Group*] *and said, where would you like to spend $500,000 on services and so 5 things came to the top*…*”. –* RCCbc Video

Autonomy as defined by the local ability to make relevant health care decisions, runs through all the data as a foundational theme in supporting system improvement.

#### Change Over Time

“Change over time” is a prominent contextual factor that underpins all the themes within the SV Project to date. One of the biggest changes over time has been the change in community population. Some remote and resource-based communities reported diminishing populations, however, this was much less common than those reporting increased population growth due to young families leaving cities to find affordable housing and retirees moving in. In addition, there is a growing tourism load in many communities. These factors, exacerbated by the expectations of care for those that have moved into the community, have impacted resources and funding for longstanding residents:

> *“*…*a lot of communities are struggling with what to do with a very quickly growing, aging population*…*we have a very strong in-migration of young families*…*” – Municipality*
>
> *“*[*Our*] *patient population has increased*… [*and the*] *infrastructure has not changed*.*” – Physicians*
>
> *“*…*communities in* [*Region X*] *have been shrinking since forestry work has moved* [*away from Region X*].*” – Municipality*

Participants emphasized how demographic and population changes have created local concerns that the community services are not adapted to the changing contexts; thereby causing issues that relate to capacity, patient access, staffing, service demands, manpower, and funding that do not meet the communities’ needs:

> *“*…*our community is growing, like our nation is growing, but the services haven’t. And so, everyone’s fighting for a doc*…*” – First Nations*
>
> *“I think we’re just lacking that vision for the hospital in what is a basic level of service to serve a growing community of 21,000 that also supports 2-3 communities north of us*.*” – Municipality*
>
> *“*…*And trying to actually keep up from a staffing perspective, from a staff retention, everything from a budget, like it’s we are playing a really hard game of catch-up because it’s growing faster than we can even account for and put in services to meet the needs. That’s what I think the biggest challenge is*…*” – Health Admin*

Rural communities are dynamic and, because of their size and isolation, particularly vulnerable to changes, which may not be easily anticipated. Change is continual and only those that have the ability to find ways to adapt are able to continue to deliver effective health services.

## Discussion

The Site Visit Project has strengths in the degree of its engagement and, after engaging with 107 rural communities and conducting 382 interviews, it has shown that it is possible to collect large volumes of data about local health care issues in a systematic and meaningful way in order to influence provincial health service changes. The fact that the Site Visits team travels to each community appears to have a strong influence on the relationships and trust experienced in the interviews. Many of the interviewees have informally commented on this fact, noting that they feel that the Site Visits team now understands their remoteness, available services, difficulties with transporting patients etc., and that they feel ‘heard’. One limitation of this project is that it was carried out in British Columbia and supported by adequate resourcing through negotiated public funds allocated through the provincial physician organization. This means that it is specific to the context of British Columbia but may have elements transferable to other settings. It would only be possible to replicate this project with sufficient funding supports.

The major themes are being identified and the analyzed data shared as specialized reports to both the micro and macro policy maker levels, connecting them in a manner that is resulting in some early systemic changes. Emergency transportation is one example (https://news.gov.bc.ca/releases/2020PREM0020-000725) where the provincial government have recently announced further rural emergency transport resources. The processes described have implications for policy makers in terms of rural health, ones that can be adapted to different contexts.

The three themes described in this article appear as patterns throughout the data set. They are interlinked and can be seen as foundational elements for effective functioning of health care services in rural communities. Good relationships between providers, health authority administration, external specialist services and community members were repeatedly identified as being responsible for high functioning, successful communities. This means that effort needs to be made to create the time and space to develop relationships and that these efforts are valued by all sectors. Part of the importance of relationships was linked to the concept of autonomy which in this sense meant the ability to make local decisions when needed. Autonomy impacted both the sense of wellbeing of the partners, but could also produce very practical, rapidly implemented changes with positive results, for example in the community of Hope (https://www.youtube.com/watch?v=49GM7-AROXM&t=1s&ab_channel=TheRCCbc). The exercise of autonomy however can be problematic if not carried out within an agreed framework that requires the limits of decision making to be set and agreed with health service administration and which recognizes historical power differences in health care (15, 24). Finally, change over time is recognized as being an important contextual factor in the provision of services to small rural communities and the resilience of these communities seems related to their ability to adapt to often unexpectedly changing circumstances. Such adaptation would appear to be easier in a context of good relationships and an agreed approach to local autonomy.

There are many examples in the literature of community engagement, though the literature does not appear to contain any examples of such widespread engagement being used to support policy change at a provincial level. The SV Project benefited from the fact that it is purely about listening. It did not promise change, but rather that the information gathered would inform change. Using Boelen’s Health Care Partners model at micro and macro levels (20), the results of the SV Project are being used to discuss contextually appropriate changes for rural health care. Having all the partners present at these discussions appears to increase the chances of producing successful and sustainable outcomes. The findings fit within the “five rules of Large System Transformation” described by Best et al (25) and illustrate that rural health care is a complex adaptive system. While this study does not attempt to explore complexity, it does offer a framework for engagement and data gathering that is sensitive to complexity and local contexts and may point to an example of the paradigm shift Greenhalgh and Papoutsi call for in their editorial on studying complexity in health services research (26).

## Limitations

Not all partner groups existed or were available to meet in some communities. The latter was rare and virtual meetings were arranged when necessary.

Because the Site Visits teams were led by experienced health care providers, a power differential existed during the interviews which may have been inhibitory, particularly when interviewing Indigenous groups.

As the interviews were led by health care providers it is possible that they may have biased the discussions.

The data collected is specific to the geography, health system and rural context of BC and may not be fully transferable to other settings.

A potential future limitation may be disengagement by the communities from further site visits if there no beneficial changes are seen to occur.

## Conclusion

By modifying Boelen’s approach to partnership in health development the SV Project has demonstrated a successful way to engage rural communities and gather extensive data that can be used to inform rural health care policy in an ongoing and contextually appropriate manner. Relationships, communication and relevant data are the cornerstones that successful sustainable change is built on.

While every rural community is different, this project elicited many common themes that have linked the health care issues in rural BC. Although early changes have already occurred, further research will be needed to determine whether the changes resulting from the SV Project are beneficial and sustainable with time.

### Funding

This project is supported by funds from the Joint Standing Committee on Rural Issues through funding negotiated through the Ministry of Health and Doctors of BC’s Physician Master Agreement. Those funds are administered by the Rural Coordination Centre of BC. There is no grant number.

### Data Sharing

Due to the confidential nature of the interview data the raw data is not publicly available. All interim reports and innovations are available through public websites and links are embedded in the body of the text.

## Author Contributions

Stuart Johnston is the project lead, has been involved in all stages of the project design, has attended visits and has helped with data analysis. He wrote the first draft of the article.

Erika Belanger is the primary analyst and developed the analytic methodology, the codebook and the initial content analysis. She has attended visits and contributed to all sections of the article.

Krystal Wong developed the Site Visits processes, attended visits, has been involved in all conversation in terms of the analysis and contributed to all sections of the article.

David Snadden developed the qualitative methodology and guided the research methodology, assisted with analysis and determining themes and contributed to all sections of the article including developing and editing the final pre-submission draft.

For researcher reflexivity statements see supplementary file “Team Reflexivity”.

## Supporting information

Supplemental file interview guide

Supplemental file code book

## Data Availability

Due to the qualitative nature of the data and the fact that the raw data contains identifying information no data is publicly available

## Acknowledgements

We would like to thank the entire RCCbc Site Visits Project team for helping with the administrative coordination and reach out to communities throughout the duration of this work. We would also like to thank Gemma McEachern for providing key supports during the initial phases of this project and Anne Lesack for assisting with the data analysis throughout this project. We would like to further extend our thanks to Dr. Martha McLeod and Jason Curran for providing us with an external review and feedback on our manuscript. Thank you to the Joint Standing Committee for funding this work and allowing us to continue this project.

Most importantly, we wish to acknowledge all our participants who chose to welcome us to their communities and allowed us to listen to their experiences and perspectives. We thank you for participating in this work and contributing your voices.

