## Supplemental file interview guide for "How can rural community-engaged health services planning affect sustainable health care system changes? - A process description and qualitative analysis of data from the Rural Coordination Centre of British Columbia’s Rural Site Visits Project"

### BC Rural Site Visits Program – Meeting Guide
#### For All Health Partners

These questions are used as a guide to facilitate our meetings <u>for all health partner groups</u> (unless specified below). Meetings are semi-structured and flexible, so if there are topics that are not covered in our questions we are still very interested in discussing them with you.

**<u>General</u>**

1. Tell us about your health care in your community.
    a. What are its unique features?
    b. What works well?
2. What are your connections like with other community members?
3. How does the community support local health care?

**<u>Innovations</u>**

1. Tell us about any initiatives do you offer that you feel are successful and why?
2. Tell us about any holistic initiatives that have been put in place that support a person's well-being spiritually, mentally, and/or physically?
3. Are there any unique solutions that you've developed?
4. What can other sites learn from you?

**<u>Access</u>**

1. Tell us about access to primary health care providers.
2. Tell us about access to specialists and other health care services.
3. How do patients get to their health care needs (ER, appointments, services, etc.)?
4. How is telehealth used in your community?
5. Are there any services at risk and why?
6. What health care services would you like to have/provide that would have the most impact for your community?

**<u>Cultural Awareness</u>**

1. With racism at the forefront of many conversations in health care, have you ever experienced or witnessed racism or other forms of discrimination/judgement when you or others are accessing/providing care?
2. What supports are there for Indigenous community members to promote cultural safety?
    *a.* Are there any supports or services in place that help promote cultural safety for staff and patients? *For example: is there a cultural space to practice ceremonies such as smudging within your hospital/clinic, is there an Indigenous liaison, are there larger spaces for families to be with the patient, etc.?*
    *b. How have these cultural safety initiatives impacted care for you/your community/your patients?*
3. **For Indigenous community members:** Tell us what would help you or a member of your community feel more culturally safe when accessing health care services?

**Pick relevant partner group:**

**For Clinicians (physicians, NPs, midwives, etc.) and Health Admin groups only: Practice Context**

1. Tell us about team-based care and/or Primary Care Networks? Describe what an ideal team-based care team would look like in your community.
2. How do health care providers in the community share the workload?
3. What workplace supports do you have (CPD, Divisions, Health Authority)?
4. How could CPD support you better?
5. Would you be interested in doing research and what supports would you need?
6. Tell us about any real-time support initiatives.
7. Tell us about any locum support in your community.

**For First Responders group only**

1. Tell us how you interact with the local health care providers?
2. Tell us about any locum support in your community.

**For Academic group only**

1. Tell us about your teaching program.
    a. How easy is it to find preceptors?
    b. How does having learners change healthcare in your community?
2. How has having an academic program in your community affected recruitment and retention?

**Recruitment and Retention**

1. How do you address recruitment of health care providers?
2. How do you retain health care providers in the community?
3. Are there any supports available for the spouses/family members of those being recruited to the community?

**Concluding Questions**

1. How has Covid-19 affected health care in your community?
2. What keeps you up at night? What is your main worry?
3. What are you proud of?
4. Have we missed anything else you would like to contribute?
5. Do you have any feedback on this process?
