## Supplemental file code book for "How can rural community-engaged health services planning affect sustainable health care system changes? - A process description and qualitative analysis of data from the Rural Coordination Centre of British Columbia’s Rural Site Visits Project"

### Site Visits Master Codebook

Developed by: Erika Belanger
Updated by: Erika Belanger + Anne Lesack on November 28 2019

#### Nodes\\Themes

Legend: Parent Nodes = Black
Child Nodes = Orange
Grandchild Nodes = Red

| Category | Description |
|---|---|
| Advocacy | Those who advocate or stand up for the health needs of the community. Can be a community member, physician, someone from municipality, or a group of individuals who the community trusts to speak on their behalf. Typically, this individual or group of people have strong interconnected ties with the community and has an in-depth understanding of an area in health care. |
| Alternative Healing | Health-related services that are already offered, or wish to be offered, outside of the traditional "western-way" of medicine and service delivery. This may include services/activities that focus on mental/spiritual/cultural health that are (or can be) practiced at an individual or group level within a community. |
| Areas of Opportunity | Areas of health care that provide an opportunity to be changed or improved upon within reason. Examples range from old & damaged waiting rooms (infrastructure) to miscommunication between two or more stake holding bodies (relationship building). |
| EMR and Information Sharing | Areas of improvement which include compatibility of electronic medical records and/or paper health records. Any other information pertaining to the improvement of information sharing, monitoring and/or access of health data is included. |
| Education and Training | Opportunities for education and/or training for health professionals and/or health partners. |
| Equipment | Equipment that needs to be replaced or updated. |
| Funding | Areas in which funding could be allocated (e.g. health, service delivery, program implementation, etc.) |
| General Safety | Situations that are placing (or potentially placing) physicians, nurses, community stakeholders, or patients at risk. Includes: occupational safety, community safety, etc. *Note: situations that appear to be putting individuals in serious and/or immediate danger should be reported to RCCbc management ASAP.* |

| | |
|---|---|
| Housing | Areas where lack of housing is identified. This includes housing for general community members, medical residents, physicians, and locums. |
| Infrastructure | Infrastructure (buildings, roads, telecommunication services), that need to be built, replaced, fixed, or upgraded. |
| Manpower & Coverage | Areas where more coverage is needed and the desire exists to have another health-professional body present. Also relates to scenarios in which individuals are feeling short-staffed and stretched too thin to be performing at an optimal work-level. |
| Policy Change | Policies, regulations, local rules processes and measures that could be changed to improve community outcomes. (may move this node in future) |
| Relationship Building | Areas that demonstrate poor communication, lack of team building or connection building etc. Scenarios where individuals feel misunderstood are also included. |
| >Collaboration | Situations in which there is a lack of collaboration or cooperation between individuals or groups on different levels in different areas. Lack of cooperative action towards a common goal. Also includes areas where collaboration can take place between two groups to better health service delivery. |
| >Communication | Areas/situations where there is a lack of information exchange and/or open communication between individuals and/or groups. |
| >Developing Trust | Participants indicate a need for increased trust or a noted lack of trust in their relationship with an individual/partner/organization/Health Authority/ group. |
| >Transparency | Participants indicate desire for more/ or indicate a lack of openness, honesty and clarity in their relationship with an individual/partner/organization/Health Authority/ group. |
| Research | Expressions from physicians, residents, or other individuals who wish to take part in research within their community. |
| Support | Areas in which direct support or additional support is requested by any health care partner in any area. |
| Understanding Awareness and Recognition | Participants express a gap in an ones own/ individuals/ groups/ HA's, etc. understanding, awareness, recognition or knowledge regarding an aspect of health service delivery, community rurality, cultural practice, etc. |
| Change Over Time | Any reported change that has occurred within a community over any given period of time. This can be a health-service related change but may also be a change in community priorities, initiatives, group beliefs, relationships, finances, etc. |

| | |
|---|---|
| Confidentiality | Thoughts, feelings, perspectives and/or scenarios related to personal and/or patient confidentiality, identity, and reputation. |
| Demographic Focuses | Health care focuses, successes, and challenges that relate to a specific demographic within a community. |
| Aging | Focuses related to aged or aging individuals within a community. |
| Families | Focuses related to families in a community. |
| Youth | Focuses related to youth in a community. |
| Discharge Conditions | Conditions that patients are discharged into. (e.g. when leaving the hospital, when leaving a doctor's appointment or health care service outside of their own community, etc.) |
| Finance | Various methods of billing, funding resources, and pay models for physicians within a community. Demonstrates the variety of financial models (both successful and inadequate) utilized within communities. |
| Billing | All information pertaining to billing clinics, physicians, and/or patients. |
| Funding | All information relating to all types of funding. |
| Pay | All information relating to physician pay (or lack thereof). This includes information on different types of pay models (e.g. FFS or APP) and the successes and challenges that are shared about pay in general. This may also include information regarding outside funding that is given to physicians for their work. |
| Future Plans | Plans, initiatives, or processes that are stated to be carried out in the future. May relate to any aspect of health care. |
| Geographic Isolation | Comments related to geographic isolation; how community members perceive their level of isolation in a community. |
| Health Authority | Any reference to interactions with a communities HA and/or to assistance, successes, challenges brought upon a community through their HA. May also include information regarding communities that declare the presence/absence of their ties with their HA. |
| Interior Health | All comments about/directly involving Interior Health. |
| First Nations Health Authority | All comments about/directly involving FNHA. |
| Fraser Health | All comments about/directly involving Fraser Health. |
| Northern Health | All comments about/directly involving Northern Health. |

| | |
|---|---|
| Vancouver Coastal Health | All comments about/directly involving Vancouver Coastal Health. |
| Vancouver Island Health | All comments about/directly involving Island Health (also known as Vancouver Island Health, VIHA). |
| Health Care Approaches | Approaches that are taken in regards to service delivery, funding, etc. that is implemented in a specific manner. |
| Bottom Up | Initiatives that are developed by people in a community, for people in that community. Decision making on program and service development, service implementation, recruitment, and/or funding, are made directly by community members, who identify what the needs are in the community. |
| Top Down | Initiatives that are developed by people that do not live within a community (i.e. those that sit in higher governing bodies), that must be followed by people living in that specific community. With this approach, community members are directed to follow decisions made by those who are removed from the community – typically for things such as service delivery, funding, recruitment, etc. |
| Siloing | Dialogue that explicitly discusses siloing. |
| Centralizing | Dialogue that explicitly discusses centralizing or centralization of health services |
| Indigenous | All information that pertains specifically to/from First Nations. |
| Alternative Healing Practices | Specific comments from First Nations around health services /practices outside of the traditional “western-way” of medicine and service delivery. This may include services/activities that focus on mental/spiritual/cultural health that are (or can be) practiced at an individual or group level within a community. |
| Connection With Others | Connections that a group of First Nations have with eachother (in their own band/community e.g. caring circle, interprofessional teams) or that they have with other members of a community. Includes their relationships with others (good or bad), their expressed desire to have relationships with certain people/groups of people and/or connections that can be improved upon. |
| Cultural Safety | Includes comments around experiences, perceptions and views of cultural safety within medical and community environments. |
| >Needed | Participants express a need for, or a lack of cultural safety within medical or community environment. Can include comments around: racism, lack of time, lack of listening, lack of cultural awareness, etc. |

| | |
|---|---|
| >Provided | Participants express situations in which culturally safe care was delivered, experienced or demonstrated in a health or community environment. |
| Culture and Identity | Comments around culture and/or identity. Also includes loss/gain of culture and/or history |
| General | This section includes all of the “Indigenous” information that was formerly under “Demographic focuses -> Indigenous” Everything that is related to First Nations specifically that does not fall under any other category under the “indigenous” node is coded here. |
| Access and Service Delivery | Health care services that are offered and/or accessed within an Indigenous community. (e.g. community nurse that works with the band, community social worker specifically for the band, etc.) |
| Trauma | Comments around impact or experience of trauma by oneself, within a community or intergenerational trauma. |
| Innovations | New or unique method, idea, product or workaround that benefits a community’s health service delivery in any way. |
| Locums | Any information regarding the ability to bring in locums into a community, how locums contribute to a community, and the ease in which a community can access locums for any given period of time. |
| New to Practice Physicians and Students | Impacts, impressions, and overall effect that new physicians and/or residents and/or students establish while practicing in a rural community; this includes comments regarding perceptions of health care providers about new to practice physicians and work style. (This node was formerly known as new grads and residents) |
| Nursing | Any items related to nursing in the context of rural health and health care delivery. |
| Patient Capacity and Attachment | Information relating to wait-times for services, family physician availability, or number of beds available within a hospital setting. Includes accounts relating to patient attachment and how patients are attached/unattached in a community. |
| Population | Health and non-health related (i.e. community events) aspects of a population that relate to a community’s population growth, recruitment, and retention. |
| Decline | References of population decline within a community. |
| Growth | References of population growth within a community. |
| Recruitment | References of recruitment into a community. Recruitment successes and challenges are included. |

| | |
|---|---|
| Relocation | References of relocation into or out of a community. Relocation successes and challenges in a community are included. |
| Retention | References of retention in a community. Retention successes and challenges are included. |
| Tourism | References of tourism in a community |
| Proposed and Potential Solutions | Initiatives that have been proposed, suggested, or are in the beginning stages of implementation for the purpose of addressing/overcoming a challenge within a community. |
| Powerful Quotes | Meaningful quotes that shed light on positive, unforeseen, or unique aspects of healthcare in a community. |
| General | General quotes as defined by the "Quotes" category description. |
| Questions | Questions that participants ask as defined by the "Quotes" category description. |
| Stories | Stories that participants share as defined by the "Quotes" category description. |
| PRA's and IMG's | Any information that relate to PRA's and/or International Medical Graduates (IMG's). |
| Programs and Networks | Information that relates to specific programs and networks and how community members find these things either beneficial/not beneficial in their community. May also include accounts where individuals note that they have not heard about a specific network/program. |
| CPD | Any comments related to continuing professional development and continuing medical education. |
| Divisions | Any comments related to divisions of family practice. Includes both positive and negative accounts surrounding divisions; interactions, assistance, and successes brought upon a community through their respective divisions group. May also include information regarding communities that declare the presence/absence of their ties with a division. |
| JSC Programs/Initatives | All program information that relates to a JSC program below. |
| >NITAOP | Any comments related to the Northern & Isolation Travel Assistance Outreach Program (NITAOP). |
| >REAP | Any comments related to the Rural Education Action Plan (REAP) |
| >REEF | Any comments related to the Rural Emergency Enhancement Fund (REEF). |

| | |
|---|---|
| >RSON | Any comments related to the Rural Surgical and Obstetrical Networks (RSON). |
| >RRP | Any comments related to the Rural Retention Program (RRP). |
| SSC Programs/Initiatives | All program information that relates to an SSC program below. |
| >Facility Engagement | Any comments related to facility engagement and/or interactions with facility engagement liaisons (FELs). |
| PCN’s | Any comments related to the Primary Care Networks (PCN’s). |
| MOCAP | Any comments related to the Medical On Call Availability Program (MOCAP). |
| RCCbc Connection Points | Areas where RCCbc staff/core members are able to connect people with eachother and/or information. Includes feedback that is received on the Site Visits Project. |
| Follow Up's | Questions that participants have that RCCbc staff can answer and follow up on; and areas in which RCCbc staff can offer connections to other individuals or advice on a given topic. |
| Project Feedback | All feedback that participants share with regards to the Site Visits Project. |
| Resource Development | Comments that are made about resource development in a community. May include how resource development has directly/indirectly affected a community (e.g. mining, LNG project, watersheds, logging, farming, ecosystem etc.) |
| Rural vs Urban Perspectives | Any comparison or contrast between a rural community and another (typically urban) community that either: (i) has more services offered and/or (ii) is a larger referral community. Note: some communities may compare themselves to a larger community that is also rural. While larger rural communities are not urban, smaller rural communities may refer to these larger rural communities as so due to the above reasons. |
| Scope of Practice & Workload | The entire role that physicians and/or other health professionals encompass as a rural health care provider. This may include general and or specific skill sets that are required from individuals in a given community. Other concepts included in this section are physician expectations (from self and others), physician wellbeing, and physician burnout (associated with heavy workloads, lack of time off, etc). |

| | |
|---|---|
| Physician Wellbeing | Any part of a rural physician's scope of practice that relates to a physicians' well-being. Includes info that may lead (or has led) to physician burn-out |
| Physician Time Off | Any part of a rural physician's scope of practice that allows/does not allow adequate time off |
| Services | Any health-related service that is at risk of becoming extinct or in need because that service is (1) currently not available in the area and (2) currently in significant demand by patients and health providers. |
| At Risk | Services at risk. |
| In Need | Services in need (general). |
| >Mental Health and Addictions | Mental health and addiction services that are needed, or accounts that describe where/why such services are needed (specific). |
| >Obs, Gyn, and Maternity | Obstetrics, Gynecology, and/or Maternity services that are needed, or accounts that describe where/why such services are needed. (specific). |
| Lost | Services that were once offered but are now obsolete. |
| Social Determinants | Measures related to socioeconomic status that affect the health status and use of health services by individuals. |
| Successful Initiatives | Initiative such as measures, models, programs, methods, or systems that have created a beneficial impact in improving the health care and/or health service delivery of a community. |
| Measures | Measures such as having enough staff, having successful community support etc. that contributes to health care and service delivery success within a community. Includes initiatives that do not fall under the "models" or "programs" category, |
| Models | Models such as funding models, clinic models, etc. that contributes to health care and service delivery success within a community. |
| Programs | Any program that has been implemented/delivered etc. that contributes to health care and service delivery success within a community. |
| Support | Supports that are essential and contribute to maintaining successful health care outcomes within a community. |
| Collaboration & Connection | Scenarios where individuals from different areas (of profession or of geographical location) connect with each other on some level (i.e. communication, decision making) to improve an aspect of health care. Included in this section are examples of individuals or groups connecting with each other in order to: a) work together towards a common goal or outcome; or b) share ideas in a collaborative manner. Relationships that have been built between two entities may also be included. |

| | |
|---|---|
| Community Support | Support that is provided by general members within a community, or by community members that work in community-focused groups such as municipality, volunteer organizations, and/or community health organizations. |
| Employee Support | Support that is provided by employees towards each other in a given setting. |
| >Culture | Successful work-cultures that employees create within their working environment. |
| >Dedication | Expressions of commitment and dedication for work, delivery of services, and or towards patients/community members within a given profession. |
| >Teamwork | Areas in which teamwork/collegiality has been highlighted/demonstrated within the workplace. |
| Telehealth | Information, including successes and challenges, relating to telehealth services. |
| Time | Situations in which time has a significant impact or is mentioned as important in a given situation (e.g. physicians expressing they need more time with their patients, etc.) |
| Transportation | All methods of transportation utilized by community members for local and long-distance transport. This section includes specific methods, thoughts, successes and challenges related to local transportation, emergency transportation, accessing areas far away (distance) and environmental factors/conditions. |
| Alberta proximity | Information relating to successes/challenges that derive from communities that are in close proximity to the Alberta border. |
| Distance | Non-emergency transportation that requires an individual to travel a distance outside of their community for health care services. Examples include: needing to travel out of town for cancer appointments/dialysis/regular GP appointments, etc. |
| Local | Non-emergency transportation that requires an individual to travel within the community for health care services. This includes information related to the availability of taxis/buses/volunteer drivers/etc within a community. |
| Emergency Transport | Successes and challenges related to emergency transportation. |
| Environmental Factors | Environmental factors that affect the ability to transport into and/or out of a community. |
| >Weather | Scenarios in which weather has impacted transportation. This includes the ability to enter/leave a community. |

| | |
|---|---|
| >Wildfires | Scenarios in which wildfires have impacted transportation. This includes the ability to enter/leave a community. |
| >Flooding | Scenarios in which flooding has impacted transportation. This includes the ability to enter/leave a community. |
| Patient Transfer Network | All information pertaining to the Patient Transfer Network (i.e. successes and challenges) |
